# Proportional Relative Hazards Model for Competing Risks Data

**DOI:** 10.1101/2020.08.27.20183244

**Authors:** Hanjie Shen, Jong H. Jeong, Loren K. Mell

**Author notes:** **Correspondence** Prof. Loren K. Mell, Department of Radiation Medicine and Applied Sciences, 3855 Health Sciences Drive, MC0843, University of California San Diego, La Jolla, CA, 92037, U.S.

## Abstract

In this article, we propose a Proportional Relative Hazards (PRH) model to differentiate subjects according to their risk for a primary event relative to competing events. The model estimates effects on the baseline ratio of the hazard for a primary event, or set of primary events, relative to the hazard for a competing event, or set of competing events (*ω*^+^ ratio). An analogous model is presented to estimate effects on the baseline ratio of the hazard for a primary event (or set of events) relative to the hazard for all events (*ω* ratio). A weighted regression method is introduced, along with practical presentation of risk-stratification using the PRH model in breast and head and neck cancer data sets.

## 2 BACKGROUND

Clinical researchers are often interested in estimating event probabilities and effects of treatments and/or covariates in populations at risk for competing events. For example, in oncology, researchers often consider various outcomes including cancer-specific mortality, specific patterns of cancer recurrence or progression (i.e., local, regional, or distant), and competing causes of mortality due to treatment or comorbid illness. Often these events are pooled together in comprising a composite end point for analysis.

A problem with applying standard models to composite endpoints is that the events comprising the endpoint are treated interchangeably, resulting in the pooling of patients at high (or low) risk for primary events with those at high (or low) risk for competing events. Most cancer staging systems, for example, assign stage based a patient’s risk of mortality regardless of cause. Asa result, such models lead to inefficient risk stratification, because primary and competing events are generally at odds with each other with respect to the benefit of a treatment: a patient at high risk of dying from cancer and low risk of dying from competing causes stands a greater chance of benefitting from cancer therapy than one with the opposite risk profile.

While previous authors have developed models applicable to competing risks settings [1, 2, 3], these models do not directly address variation in the relative probability of primary vs. competing events, even though this ratio is a principal driver of the overall treatment effect. For example, previously, Mell and Jeong (2010) [5] showed that in competing risks settings, assuming proportional hazards, the overall hazard ratio (Θ) can be re-expressed as:

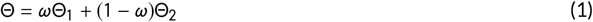

where Θ_1_ is the hazard ratio specific to the primary event(s) of interest, Θ_2_ is the hazard ratio specific to the competing event(s), and *ω* is the ratio of the baseline hazard for the primary event(s) to the baseline hazard for all events. It follows that, in addition to the intrinsic effects of therapy on both primary and competing events, the value of *ω* is a primary determinant of a treatment’s benefit. Recent research has thus focused on developing models to stratify patients [10,11] and predict treatment effects [8,9] according to the *ω* ratio.

Notably, Nicolaie et al. [4] proposed vertical models as an approach to analyze competing risks data with missing failure types. However, when the causes of failure are largely known, a practical and critical question is to identify an optimal method to select patients for more vs. less intensive therapy. The purpose of this study is therefore to present a Proportional Relative Hazards (PRH) model, with the principal objective to differentiate subjects according to their risk for a primary event relative to competing events. We use simulated data to examine several candidate methods to estimate the w+ and w functions, and briefly examine extensions of the PRH model to subdistribution hazards. Finally, we introduce a framework called weighted PRH regression, with practical application to both population-based and clinical trial data sets.

## 3 METHODS

### 3.1 Baseline *ω*^+^(*t*) and *ω*(*t*) Functions

In the PRH model, we first seek to estimate the baseline function *ω*^+^(*t*), which represents the risk for a primary event (or set of events) of interest relative to the risk for a competing event (or set of competing events). Analogously, we will also estimate the baseline function *ω*(*t*), which represents the risk for a primary event (or set of events) of interest relative to the risk for both primary and competing events. Here we compare several candidate methods to estimate these functions: (1) a “natural” or naïve method that ignores risk sets, (2) a method based on Nelson-Aalen estimator for cumulative cause-specific hazards, and (3) a method based on cumulative incidence functions.

### 3.2 Natural Estimation of Baseline *ω*^+^(*t*) and *ω*(*t*)

Let 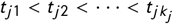 denote *k_j_* distinct failure times for type *j* and *d_ji_* denote the number of death due to cause *j* at time *t_ji_*. Hence,

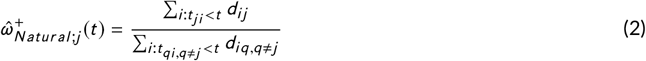

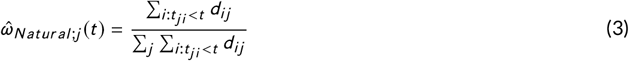

### 3.3 Nelson-Aalen Estimation of Baseline *ω^+^*(*t*) and *ω*(*t*)

Let 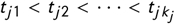 denote *k_j_* distinct failure times for type *j*, *n_ji_*, denote the number of subjects at risk due just before *t_ji_*, and *d_ji_* denote the number of death due to cause *j* at time *t_ji_*. Hence,

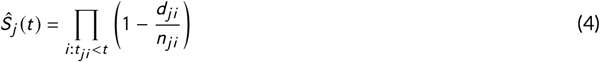

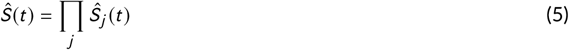

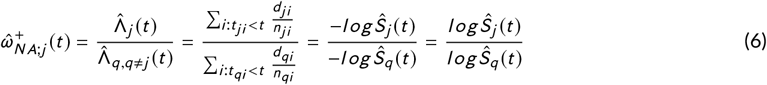

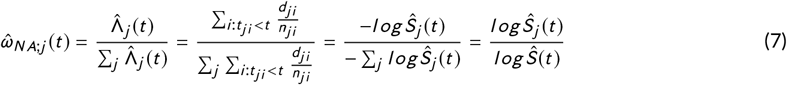

### 3.4 Cumulative Incidence Estimation of Baseline *ω*^+^(*t*) and *ω*(*t*)

Let *t*_1_ *< t*_2_ *< … < t_k_* denote *k* distinct failure times, *d_ij_* denote the number of death due to cause *j* at time *t_i_*, *n_i_* denote the total number of death at time *t_i_*, and *Ŝ* (*t_i_*) is the standard Kaplan-Meier estimator of survival at time *t_i_*. Hence,

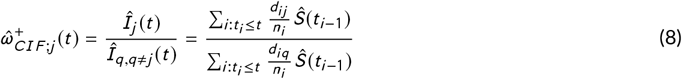

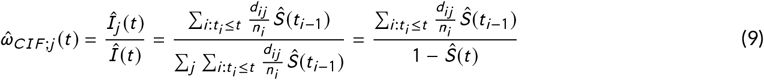

### 3.5 Proportional Relative Hazards Regression Model

In the PRH regression model, we seek to estimate effects on the *ω*^+^(*t*) and *ω*(*t*) function, rather than the hazard or subdistribution hazard. Therefore, for mutually exclusive events of type *k*, we propose the following proportional relative hazards (or subdistribution hazards) model:

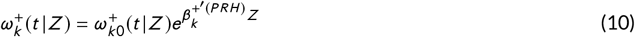

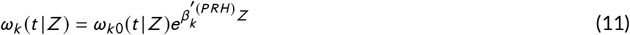

where 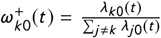 and 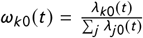. Here *.λ_k_*_0_(*t*) is the baseline hazard (or subdistribution hazard) for an event of type *k*, ∑*_j≠k_ λ_j_*_0_(*t*) is the baseline cause-specific hazard for the set of events competing with event type *k*, ∑*_j_ λ_j_*_0_(*t*) is the baseline cause-specific hazard for all events, *Z* is a vector of covariates, 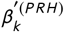 and 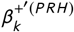 are the vectors of effects (coefficients) on the covariates. For subdistribution hazards, ∑*_j≠k_ λ_j_*_0_(*t*) does not have the same interpretation as for cause-specific hazards, and instead represents the sum of the individual subdistribution hazards for events competing with event *k*, while ∑*_j_ λ_j_*_0_(*t*) represents the sum of the individual subdistribution hazards for all events including type *k*. The PRH regression estimators then are defined as:

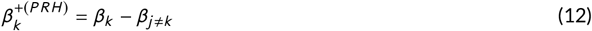

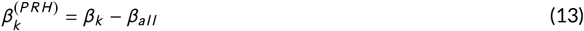

where *β_k_*, *β_j≠k_* and *β_all_* represent effects on the baseline hazard (or sub-distribution hazard) for event type *k*, competing events, and all events respectively, from the Cox Proportional Hazard model or Fine-Gray Model.

We use 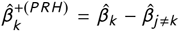 as the estimator of 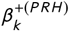 and 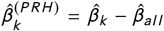 as the estimator of 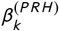 Clearly, 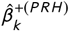 and 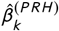 are unbiased as the partial likelihood estimators are unbiased. An R package (gcerisk) [6] was developed to facilitate this type of analysis.

Next, we want to estimate the predicted values of 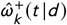 and 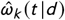 for an individual with given data vector *d*:

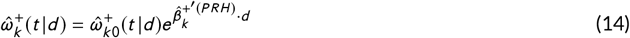

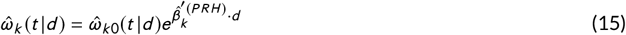

However, since the baseline *ω* function is bounded between 0 to 1, equation (15) could result in non-sensical values for *w_k_* (*t* |d). Therefore, we can approach this in a different way, by estimating the predicted value of 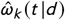 as:

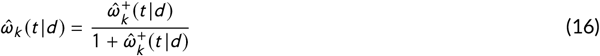

Note that while 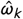 ranges from 0 to 1 inclusive, 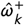 can range from −∞ to ∞. For *k* = 2, a value of 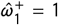 means the hazard for event type 1 equals the hazard for event type 2, and therefore 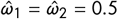.

### 3.6 Variance of PRH Estimators

Consider the situation with two type of events (primary event and competing event), we have the following two proportional hazards models:

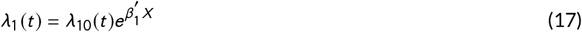

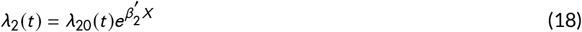

Hence, we have *β^+^*^(^*^PRH^*^)^ = *β*_1_ −*β*_2_. Consider the Cox regression model with covariates *x*,*δx* stratifying by event type, *δ* = 1 or 0 (1 is primary event and 0 is competing event). Hence, the partial likelihood function is

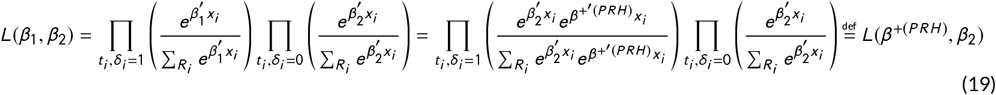

where, *R_i_* is the risk set at failure time *t_i_*. A main assumption for this approach is that we do not know the relationship between two baseline hazard functions *λ*_01_ and *λ*_02_. And then, the maximum likelihood estimator 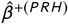 and 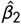 can be found by solving the derivative of log partial likelihood function. Further, the standard likelihood theory can show that

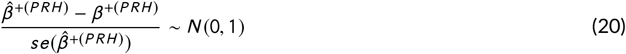

The variance of 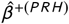 can be estimated by inverting the second derivative of the partial likelihood, which is 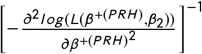, replacing *β*^+(PRH)^ and *β*_2_ with 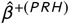 and 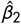. The variance of 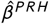 can be estimated similarly.

### 3.7 Weighted Proportional Relative Hazards Regression Model

A more advanced and tunable method is proposed as weighted PRH Regression. In this method, the risk-stratification is based on weighted risk scores 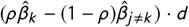 for each individual from low to high, where *ρ* ∈ [0,1] is the weight parameter to measure the relative importance of the primary event compared to the competing event and *d* is the data vector. The *ρ* parameter is treated as fixed and could be chosen to emphasize the stratification according to a specific event; in unweighted PRH regression this parameter is omitted.

## 4 SIMULATION AND RESULTS

### 4.1 Simulation Study for Comparison among Estimation from Natural, Nelson-Aalen and Cumulative Incidence Function

With the assumptions of constant hazard function, we assume that time *T ~ Weibull* (*b, p*) with probability density function

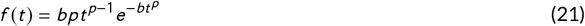

where *b >* 0 and *p >* 0. Hence, the hazard function is given by

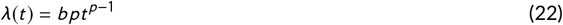

Further, we have the survival function:

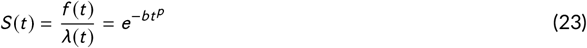

where *S*(*t*) is survival function, *b* is scale variable for *Weibull* distribution, *p* is shape parameter for *Weibull* distribution and *t* is time. Hence, we will have

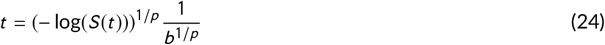

Further, by using the probability integral transformation with fixed baseline hazard as *b* for primary event (*λ*_10_) and competing event (*λ*_20_), we can generate the time vector from *Weibull* distribution with shape parameter *p* and scale variable *b* = *λ*_10_ + *λ*_20_. Then we assign the event times to primary event with probability of 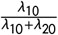, and the other will be the competing event [7]. One third of the events were treated as censored.

Based on the procedure above, we generated 5000 competing risk datasets with 200 observations for each dataset based on fixed baseline *ω* values of 0.67 (*λ*_10_ = 0.5, *λ*_20_ *=* 0.25) and 0.8 (*λ*_10_ = 1, *λ*_20_ *=* 0.25) (i.e., baseline *ω^+^* values of 2 and 4), and shape parameter *p* = 1 as the hazard is constant. The comparison between the sampling distributions of three type of estimation is given in the density plots (Figures 1 and 2). Figure 1 shows that the sampling distribution of *ω* appears approximately normal for each of the candidate estimators, suggesting they are consistent. Figure 2 shows the estimators for *ω^+^* are right skewed.

#### 4.1.1 Sampling Distribution of Baseline *ω*(*t*) Estimation

**FIGURE 1.**
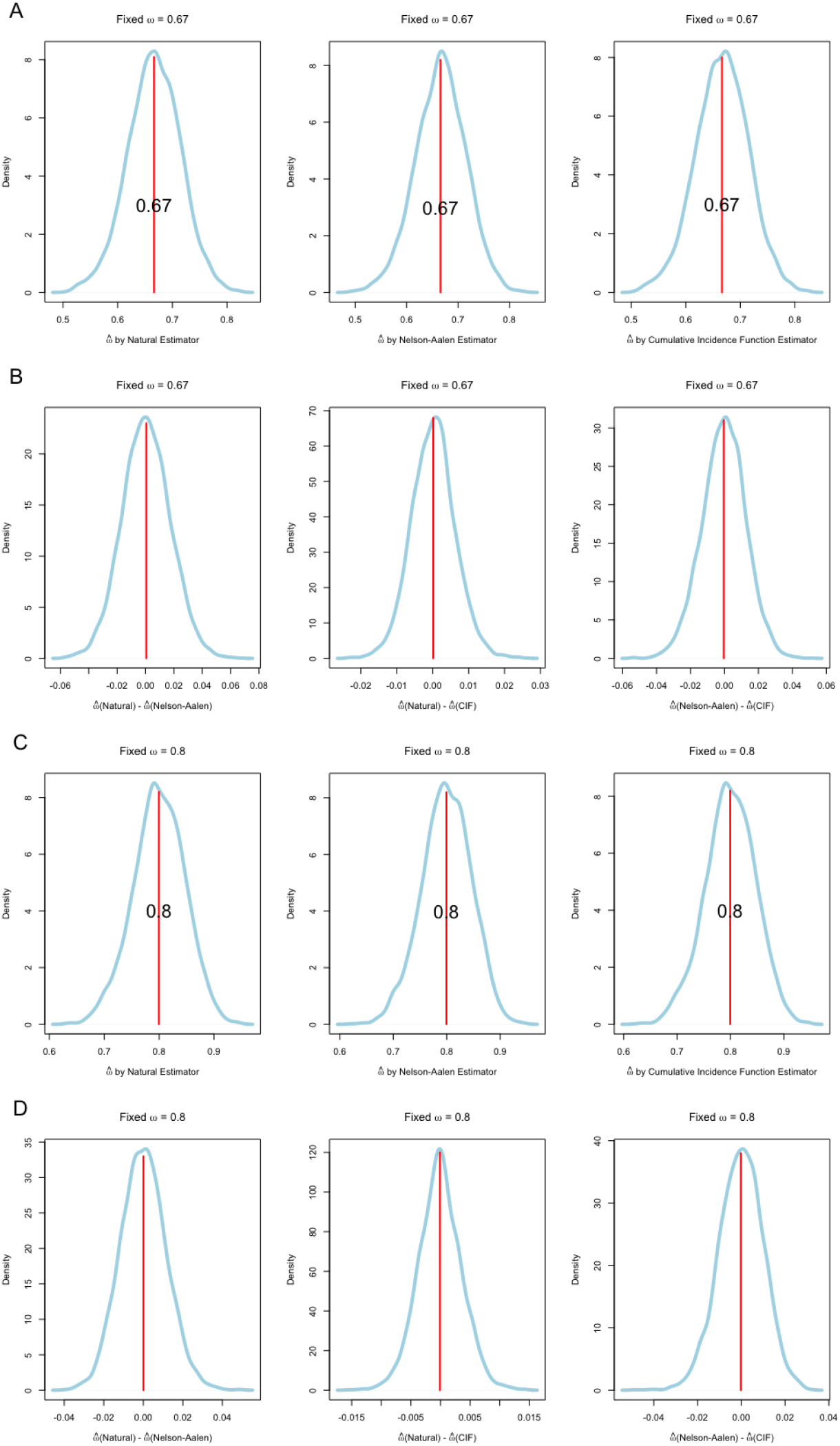
Density Plots of 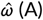 (A) Sampling Distribution of 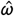 from Natural, Nelson-Aalen and Cumulative Incidence Estimation (*ω* = 0.67). (B) Sampling Distribution of Difference of 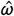 among Natural, Nelson-Aalen and Cumulative Incidence Estimation (*ω* = 0.67). (C) Sampling Distribution of 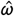 from Natural, Nelson-Aalen and Cumulative Incidence Estimation (*ω* = 0.8). (D) Sampling Distribution of Difference of 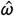 among Natural, Nelson-Aalen and Cumulative Incidence Estimation (*ω* = 0.8).

#### 4.1.2 Sampling Distribution of Baseline *ω*^+^(*t*) Estimation

**FIGURE 2.**
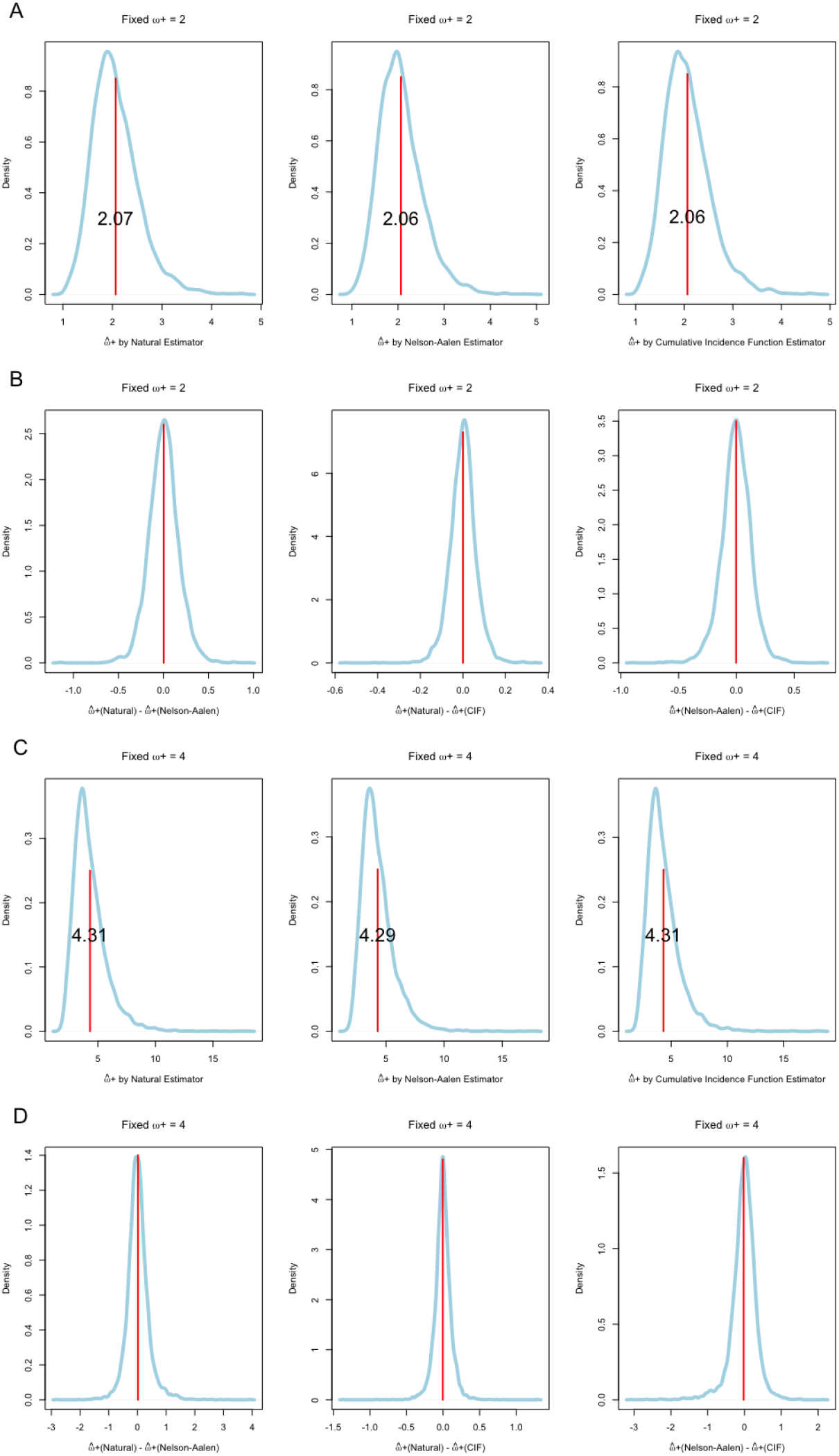
Density Plots of 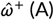 (A) Sampling Distribution of 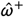 from Natural, Nelson-Aalen and Cumulative Incidence Estimation (*ω^+^ =* 2). (B) Sampling Distribution of Difference of 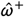 among Natural, Nelson-Aalen and Cumulative Incidence Estimation (*ω^+^* = 2). (C) Sampling Distribution of 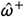 from Natural, Nelson-Aalen and Cumulative Incidence Estimation (*ω^+^ =* 4). (D) Sampling Distribution of Difference of 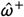 among Natural, Nelson-Aalen and Cumulative Incidence Estimation (*ω^+^ =* 4).

### 4.2 Effectiveness of Risk-Stratification using the PRH Model

First, using the SEER-Medicare database, we identified 22,929 patients with non-metastatic primary breast cancer diagnosed from 2004 to 2009. In this demonstration, we consider a primary event to be death from either non-cancer causes or due to a secondary malignancy, whereas the competing event is defined as death due to breast cancer. Surviving patients were censored at last follow-up. Covariates included in this model are age at diagnosis (continuous), sex, race (black vs. white/other), marital status (married vs. unmarried), median household income (lower vs. higher than mean), stage, grade, and modified Charlson comorbidity index, region (West vs. East vs. Midwest vs. South), teaching hospital (yes vs. no), tumor size (< 2 cm vs. ≥ 2 cm to < 5 cm vs. ≥ 5 cm vs. size unknown), estrogen receptor and/or progesterone receptor status (positive vs. negative vs. unknown), sentinel lymph node biopsy (yes vs. no), axillary lymph node examination (yes vs. no), tumor laterality (left vs. right), surgery (mastectomy vs. lumpectomy), chemotherapy (yes vs. no), and RT technique (hypofractionated vs. conventional). The PRH risk strata were defined according to quantiles of the distribution of predicted *ω* values, which were calculated from the equation (14) and (16). The unweighted PRH model is applied in order to stratify patients according to cumulative incidences and cumulative cause-specific hazards of cancer mortality vs. competing mortality within each risk group based on both the Cox model (Figure 3 A-B) and the Fine-Gray model (Figure 3 C-D).

**FIGURE 3.**
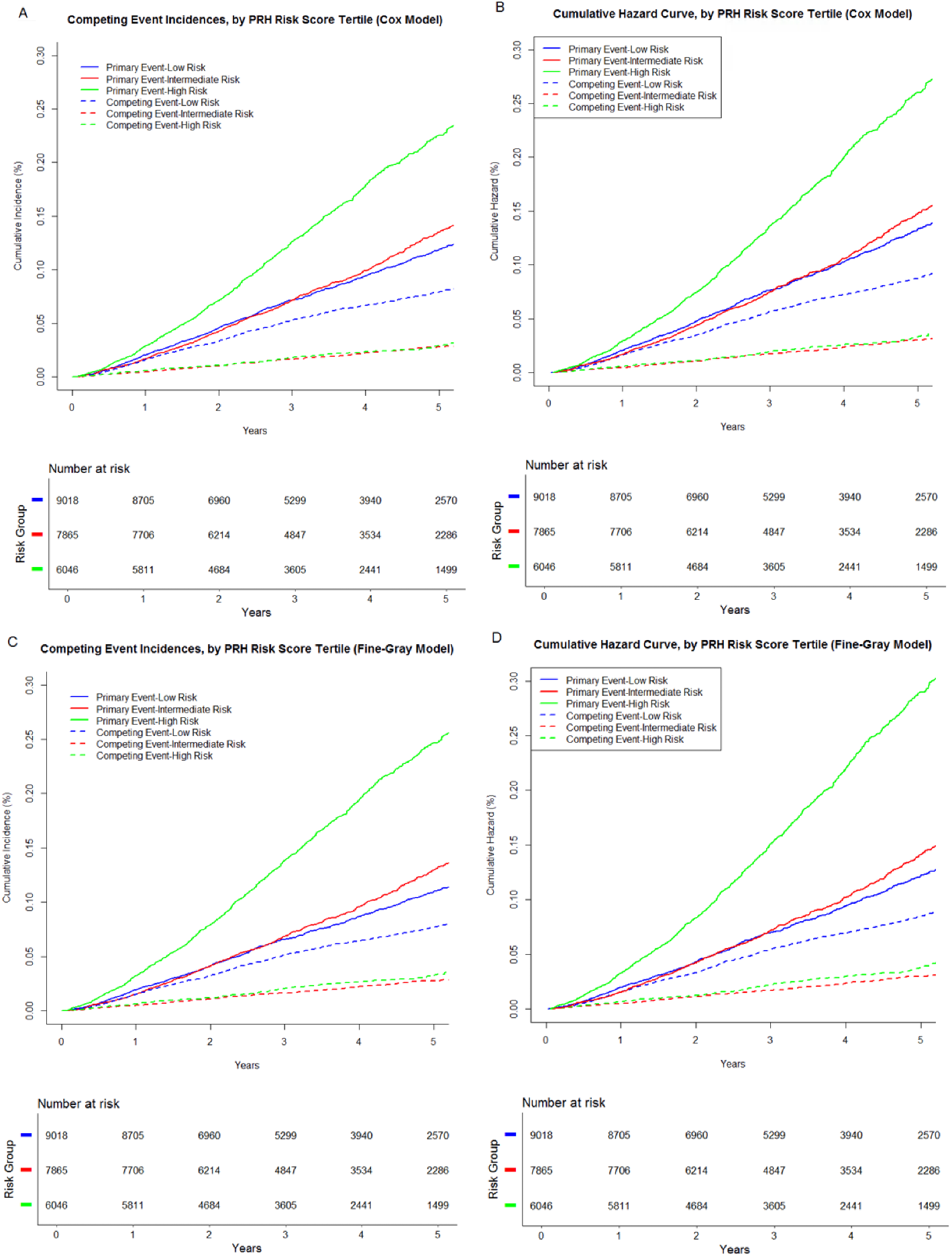
Risk-Stratification According to Unweighted PRH Models. (A) Cumulative Incidence Curves from PRH Regression based on Cause-Specific Hazards (Cox Model) (B) Cumulative Hazard Curves from PRH Regression based on Cause-Specific Hazards (Cox Model) (C) Cumulative Incidence Curves from PRH Regression based on Subdistribution Hazards (Fine-Gray Model) (D) Cumulative Hazard Curves from PRH Regression based on Subdistribution Hazards (Fine-Gray Model)

Secondly, we applied weighted PRH regression to a database of 2,688 previously untreated locoregionally advanced head and neck cancer patients undergoing treatment with radiation therapy with or without concurrent systemic therapy on one of three randomized trials. The first instance of cancer recurrence or progression was defined as the primary event, while death from any cause in the absence of a recurrence/progression event was defined as the competing event. Covariates included in this model are age (per 10 years; continuous), ECOG performance status (0 vs. 1-2), body mass index (< 20 *kg/m*^2^ vs. ≥ 20 *kg/m*^2^), oral cavity site, N stage (0 vs. 1-3), and P16 status (positive vs. negative). The calculation of PRH risk strata here is same with analysis for breast cancer clinical trials. The weight parameter *ρ* displays changes in the relative cumulative incidence of events as a function of *ρ*. Figure 4 shows the cumulative incidences of cancer recurrence vs. competing mortality within each risk group at weight parameter *ρ =* 0.1,0.5 and 0.9.

**FIGURE 4.**
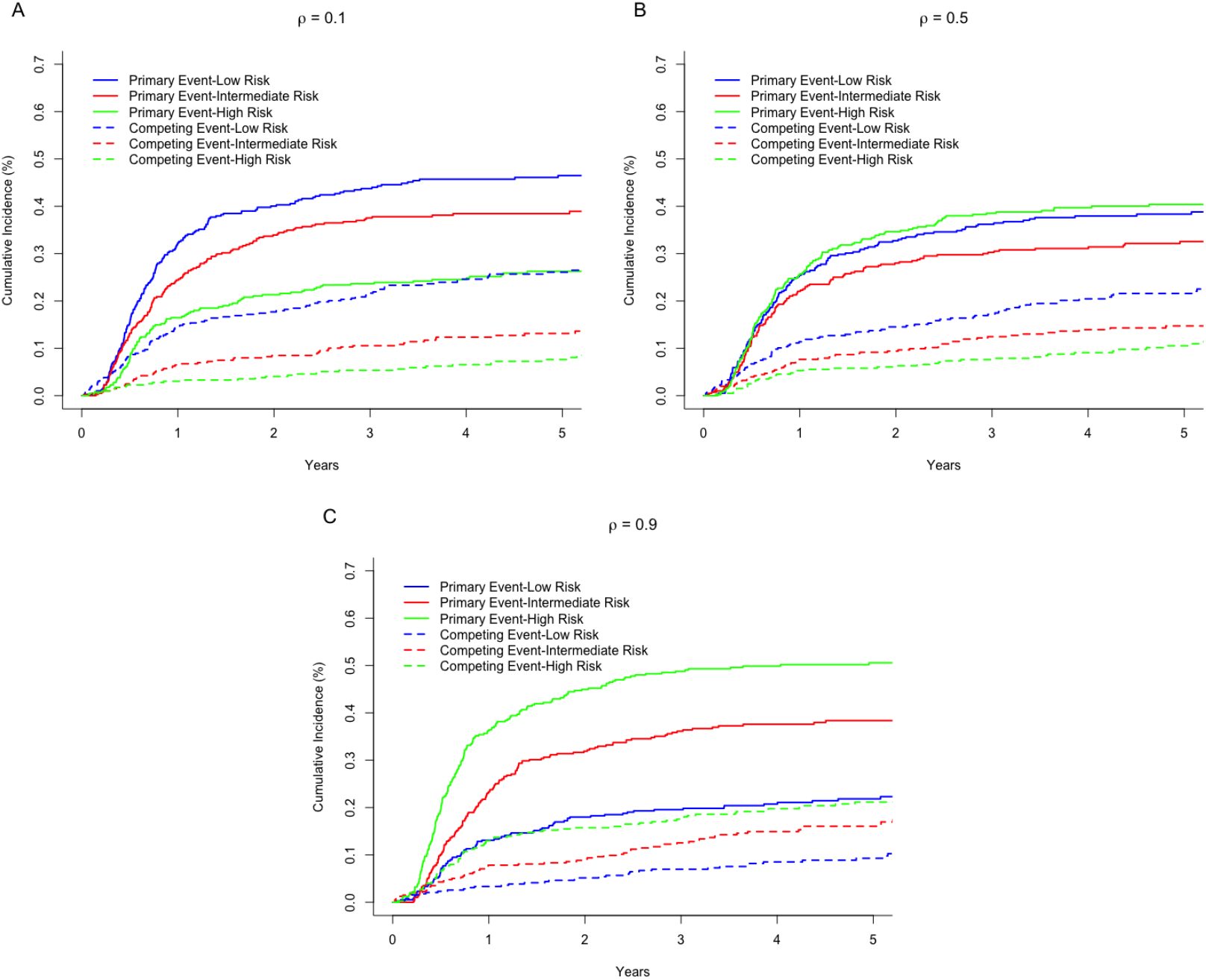
Risk-Stratification According to a Weighted PRH Model based on Cause-Specific Hazards. (A) Cumulative Incidence Curves with Weight Parameter *ρ* = 0.1 (B) Cumulative Incidence Curves with Weight Parameter *ρ =* 0.5 (C) Cumulative Incidence Curves with Weight Parameter *ρ* = 0.9

#### 4.2.1 PRH Risk-Stratification Based on Cox and Fine-Gray Regression

Unweighted PRH regression results show the ability of the model to separate samples according to patients with a high (or low) cumulative hazard or cumulative incidence of an event of interest (in this case, non-cancer mortality events) relative to a competing event (in this case, breast cancer mortality). This is because PRH models are designed to optimize the ratio of competing events in order to favor a particular event of interest. The relative probability of primary vs. competing events was increased in the highest risk strata predicted by the PRH model (higher *ω*^+^ and *ω* ratio), while the converse was true in the lowest risk strata (lower *ω*^+^ and *ω* ratio). The intermediate risk group demonstrates *ω*^+^ and *ω* ratios that are between the low and high risk strata. Note that PRH methods based on cause-specific hazards (i.e., Cox model) and subdistribution hazards (i.e., Fine-Gray model) were both able to separate the sample according to the ratio of either cumulative hazards or cumulative incidences within risk strata.

#### 4.2.2 Risk-Stratification for Patients by Weighted PRH Model with Different Weight Parameter *ρ*

Weighted PRH regression results demonstrate the ability to separate samples according to varying *ω* and *ω*^+^ ratios. In contrast to unweighted PRH regression (also called generalized competing event (GCE) regression), which has been used in several prior clinical studies to optimize risk-stratification and study effects of intensive therapy [10,11], a novel weighted PRH regression model is introduced in this paper to further adjust risk-stratification using a fixed parameter. The optimal choice of and rationale for selecting the weight parameter could be considered in further studies, with one idea to use the baseline estimate of w to weight the risk stratification.

## 5 DISCUSSION

In this analysis, we discussed a PRH modeling approach and regression method, optimally designed to separate patients according to their relative hazards for primary vs. competing events. This approach has broad utility, with the potential to improve risk-stratification of cancer patients and other subjects at high risk for competing events. When considering the effectiveness of intensive treatments, for example, it is preferable to optimize risk groups according to their predicted *ω* or *ω*^+^ value, rather than their hazard for an individual event or composite event. Our results indicate that several methods to estimate *ω* are consistent, while the *ω*^+^ estimators are right skewed. There were small differences in the mean values among the three estimators of ω. Further research is needed to explore the theoretical distribution of *ω* and *ω*^+^, to prove consistency and mathematical equivalence of the proposed estimators, and to examine analogues of *ω* and *ω*^+^ for subdistribution hazards.

Standard models used in competing risks settings are based on cause-specific hazard and/or cumulative incidence functions (CIF). Gray (1988) [1] proposed a non-parametric test to compare two or more CIFs, while Lunn and McNeil (1995) [2] applied the Cox proportional hazards (PH) regression model to competing risks based on cause-specific hazard function. Further, Fine and Gray (1999) [3] proposed a proportional hazards approach for modeling the CIF with covariates, treating the CIF curve as a sub-distribution function. Cox PH and Fine-Gray models are widely used in clinical research to analyze both composite end points and individual events of interest. However, a major limitation of these models is their inefficient risk stratification, with respect to selecting patients most likely to benefit from intensive therapy.

Additional research in the field of competing risks has focused on modeling the cumulative incidence of an event of interest in the presence of competing risks [12] and interpretation of commonly used competing risks regression models [13]. Furthermore, methods for sample size calculation [14] and analysis of clustered data [15] have been developed for competing risks research. However, other researchers have also noted the problem of inefficient stratification, which in particular impacts power estimates; see for example, Gomez and Lagakos [16].

In relation to previous work in competing risks theory, the PRH model presents a simple approach to risk stratification that could help medical researchers obtain direct information regarding effects on the ratio of primary and competing events to one another. Furthermore, better risk-stratification for the purpose of identifying candidates for intensive therapy may be found using this approach [8,9].

## Data Availability

Clinical trial data available by request from NRG Oncology. Epidemiologic data available from SEER-Medicare. All other data available upon request from the corresponding author.

## Acknowledgements

The authors would like to thank Dr. James Murphy for providing breast cancer data and the NRG Oncology Group for providing head and neck cancer data for the analysis.

## Data Availability

The data that support the findings of this study are available from the SEER-Medicare Registry and from the NRG Oncology Cooperative Group. Note that restrictions apply to the availability of these data, which were used under license for the current study, and so are not publicly available. Data are, however, available from the authors upon reasonable request and with permission from the SEER-Medicare Registry and/or the NRG Oncology Cooperative Group, as applicable.

## REFERENCES

[1] Gray, Robert J. A class of K-sample tests for comparing the cumulative incidence of a competing risk. The Annals of statistics (1988): 1141–1154.

[2] Lunn, Mary, and Don McNeil. Applying Cox regression to competing risks. Biometrics (1995): 524–532.

[3] Fine, Jason P., and Robert J. Gray. A proportional hazards model for the subdistribution of a competing risk. Journal of the American statistical association 94. 446 (1999): 496–509.

[4] Nicolaie, M. A., H. C. van Houwelingen, and H. Putter. Vertical modelling: Analysis of competing risks data with missing causes of failure. Statistical methods in medical research 24.6 (2015): 891–908.

[5] Mell, Loren K., and Jong-Hyeon Jeong. Pitfalls of using composite primary end points in the presence of competing risks. Journal of Clinical Oncology 28.28 (2010): 4297–4299.

[6] Shen, H., Carmona, R., Mell, L. K., (2018). Package ‘gcerisk’. https://cran.r-project.org/

[7] Beyersmann, J., Latouche, A., Buchholz, A., and Schumacher, M. (2009). Simulating competing risks data in survival analysis. Statistics in medicine 28(6), 956–971.

[8] Mell LK, Shen H, Nguyen-Tan PF, Nguyen-Tân PF, Rosenthal DI, Zakeri K, Frank SJ, Schiff PB, Trotti AM III, Bonner JA, Jones CU, Yom SS, Thorstad WL, Wong SJ, Shenouda G, Ridge JA, Zhang Q, Le QT. (2017). Generalized Competing Event Regression to Stratify Head and Neck Cancer Patients: Secondary Analysis of NRG Oncology RTOG 9003, 0129, and 0522 (abstr.) Int J Radiat Oncol Biol Phys. (2017) 99(2):S236–7.

[9] Zakeri K, Rotolo F, Lacas B, Vitzthum LK, Le QT, Gregoire V, et al. Predictor of Effectiveness of Treatment Intensification on Overall Survival in Head and Neck Cancer (HNC). Ann Oncol (abstr.) 2018; 29 (suppl 8): viii372–viii399.

[10] Carmona R, Gulaya S, Murphy JD, et al. Validated competing event model for the stage I-II endometrial cancer population. Int J Radiat Oncol Biol Phys 2014;89:888–98.

[11] Carmona R, Zakeri K, Green G, et al. Improved method to stratify elderly patients with cancer at risk for competing events. J Clin Oncol. 2016;34:1270–7.

[12] Kim, Haesook T. Cumulative incidence in competing risks data and competing risks regression analysis. Clinical Cancer Research 13.2 (2007): 559–565.

[13] Dignam, James J., Qian Zhang, and Maria N. Kocherginsky. The use and interpretation of competing risks regression models. Clinical Cancer Research (2012): clincanres-2097.

[14] Latouche, A., and R. Porcher. Sample size calculations in the presence of competing risks. Statistics in medicine 26.30 (2007): 5370–5380.

[15] Zhou, Bingqing, et al. Competing risks regression for clustered data. Biostatistics 13. 3 (2012): 371–383.

[16] Gómez G, Lagakos SW. Statistical considerations when using a composite endpoint for comparing treatment groups. Stat Med. 2013 Feb 28;32(5):719–38. doi:10.1002/sim.5547. Epub 2012 Aug 1. PMID: 22855368

